# Early prediction of skeletal muscle loss using longitudinal clinical data in patients with gastric cancer after radical gastrectomy and adjuvant chemotherapy: a retrospective cohort study

**DOI:** 10.64898/2026.04.28.26351920

**Authors:** Hanzheng Wang, Kun Ma, Jiezhi Lin, Juntao Zhu, Mengchen Sun, Shubin Liang, Bin Yang, Linsong Mu, Zhongchuan Lv

**Author notes:** **Correspondence** Bin Yang, Address: Department of Gastrointestinal Surgery, The Affiliated Yantai Yuhuangding Hospital of Qingdao University, NO. 20 East Yuhuangding Road, Yantai, 264000, Shandong, China., Linsong Mu, Address: Department of general surgery, The Affiliated Yantai Yuhuangding Hospital of Qingdao University, NO. 20 East Yuhuangding Road, Yantai, 264000, Shandong, China., Zhongchuan Lv, Address: Department of general surgery, The Affiliated Yantai Yuhuangding Hospital of Qingdao University, NO. 20 East Yuhuangding Road, Yantai, 264000, Shandong, China.

## Abstract

Gastric cancer patients frequently experience skeletal muscle loss during the perioperative and adjuvant treatment period, which has been associated with poorer treatment tolerance and adverse clinical outcomes. Early identification of patients at high risk of skeletal muscle loss may allow timely supportive intervention, but repeated computed tomography assessment is not always practical in routine care. This study aimed to develop an interpretable machine learning model based on routinely available clinical data for early prediction of significant skeletal muscle loss in patients with gastric cancer.

This single-center retrospective study screened 362 patients who underwent radical gastrectomy followed by adjuvant chemotherapy, of whom 292 were finally included. Significant skeletal muscle loss was defined as a decrease of at least 5% in skeletal muscle index between the baseline scan performed before surgery and the follow-up scan obtained 3 months after initiation of adjuvant chemotherapy. Candidate predictors included demographic, clinicopathological, laboratory, tumor marker, and inflammatory or nutritional variables, together with their early postoperative dynamic changes. Six machine learning models were developed and compared.

Among the evaluated models, the multilayer perception showed the best overall performance in the validation set, with an area under the receiver operating characteristic curve of 0.757 and an area under the precision-recall curve of 0.745. At the selected decision threshold of 0.45, this model achieved an accuracy of 0.693, a recall of 0.833, and a specificity of 0.525. Compared with the model using baseline variables alone, the model incorporating longitudinal dynamic features showed better overall performance. Model interpretation suggested that prediction of skeletal muscle loss was mainly related to nutritional reserve, operation-related burden, and inflammatory or metabolic status.

These findings indicate that routinely available preoperative and early postoperative clinical data can support early prediction of subsequent skeletal muscle loss in gastric cancer. This approach may help identify high-risk patients earlier and facilitate individualized nutritional support and supportive care during treatment.

## Introduction

Gastric cancer remains one of the most common and deadly malignancies worldwide[1]. For patients with resectable locally advanced disease, radical surgery followed by adjuvant therapy remains the standard treatment strategy[2]. However, skeletal muscle loss frequently occurs during the perioperative period and subsequent adjuvant treatment, and has been associated with reduced treatment tolerance, increased complications, and poor long-term outcomes[3–5]. Therefore, early identification of patients at high risk of skeletal muscle loss, before overt muscle depletion develops, is of clear clinical importance.

Computed tomography (CT)-based assessment of skeletal muscle area or skeletal muscle index (SMI) at the third lumbar vertebral (L3) level is currently regarded as the gold standard for evaluating skeletal muscle status[6–8]. Nevertheless, this method is mainly suited for objective assessment and retrospective confirmation of skeletal muscle loss, rather than early risk prediction in routine practice. Because it depends on repeated imaging examinations before and after treatment, it is relatively cumbersome and less practical for continuous monitoring. More importantly, once substantial skeletal muscle loss is detected on imaging, the optimal window for intervention may already have passed. Thus, early identification of patients at risk of subsequent significant skeletal muscle loss using routinely available clinical information remains an important unmet clinical need[9].

Routine clinical variables and their longitudinal changes may offer a practical alternative for early risk stratification[10, 11]. Inflammatory and nutritional indicators, such as the neutrophil-to-lymphocyte ratio (NLR) and leukocyte-to-lymphocyte ratio (LLR), may reflect systemic inflammation, metabolic stress, and nutritional reserve[12, 13]. However, these variables are often interrelated and may show complex nonlinear associations with skeletal muscle loss, limiting the performance of conventional statistical models. Machine learning (ML) methods are well suited to capture such complex patterns, although their limited interpretability has hindered clinical application[14].

In this study, we developed an interpretable ML model based on longitudinal clinical data to enable early prediction of significant skeletal muscle loss during treatment in patients with gastric cancer. We further applied Shapley additive explanations (SHAP) to clarify the key factors driving model predictions. Our aim was to establish an early risk assessment tool that does not rely on repeated body composition measurements, while retaining both predictive ability and interpretability, thereby supporting more precise nutritional intervention and individualized clinical management.

## Methods

### Study population

This was a single-center retrospective cohort study aimed at developing and evaluating machine learning models for predicting the risk of skeletal muscle loss during treatment in patients with gastric cancer. The study was approved by the Ethics Committee of Yantai Yuhuangding Hospital. Given the minimal-risk retrospective design, the requirement for informed consent was waived.

We retrospectively enrolled patients with gastric cancer who underwent radical gastrectomy followed by adjuvant therapy at Yantai Yuhuangding Hospital between January 2018 and December 2023. The inclusion criteria were as follows: (1) availability of a baseline CT scan performed within 2 weeks before surgery; (2) availability of early postoperative clinical and laboratory data at 1 month after surgery; (3) availability of a follow-up CT scan obtained 3 months after initiation of adjuvant therapy; and (4) complete clinical data at all required time points for model development and outcome assessment. The exclusion criteria were as follows: (1) receipt of neoadjuvant therapy before surgery; (2) no adjuvant chemotherapy after surgery; (3) lack of follow-up data; (4) CT images of insufficient quality for analysis; and (5) missing key laboratory data. Among 362 initially screened patients, 297 were finally included in the study.

Candidate variables were prespecified based on prior literature, clinical relevance, and data availability[10, 15]. These variables included demographic characteristics, tumor-related features, laboratory and tumor marker variables, and inflammatory/nutritional indicators. In addition, to capture early changes in systemic status, longitudinal change features from baseline to 1 month after surgery were further derived. Compared with single time-point measurements, these longitudinal features may better reflect the dynamic evolution of patients’ systemic condition during early treatment and thus provide more informative signals for subsequent skeletal muscle loss. Missing data were handled using multiple imputation. Before model development, continuous variables were standardized using z-score normalization to reduce scale differences and improve model stability.

### Outcome variable

CT images from all included patients were analyzed using Comp2Comp, an open-source deep learning toolkit for automated and standardized body composition assessment[16]. Developed by the Stanford Medicine Imaging and Machine Intelligence Laboratory and trained on diverse datasets, Comp2Comp enables robust automated segmentation across the L2–L4 vertebral levels. All automated segmentation results were independently reviewed by two radiologists to ensure quality control.

The outcome of interest was significant skeletal muscle loss during treatment. Specifically, changes in SMI were calculated using the baseline CT scan obtained within 2 weeks before surgery and the follow-up CT scan acquired 3 months after initiation of adjuvant chemotherapy, which approximately corresponded to 4 months after surgery. Skeletal muscle cross-sectional area was automatically segmented at the L3 level using Comp2Comp and normalized by height to derive SMI. The percentage change in SMI was calculated as follows: [(follow-up SMI − baseline SMI) / baseline SMI] × 100%. Based on a commonly used clinical threshold and our previous study, patients with an SMI decrease of ≥5% were classified into the skeletal muscle loss group, whereas the remaining patients were classified into the skeletal muscle maintenance group[10].

In addition, the NLR, LLR, and platelet-to-lymphocyte ratio (PLR) were calculated. The prognostic nutritional index (PNI) was defined as the albumin level (g/L) plus five times the absolute lymphocyte count. Furthermore, the systemic immune-inflammation index (SII) was calculated by multiplying the absolute platelet count by the NLR[17].

### Threshold selection

To support potential clinical application, we systematically assessed model performance across different decision thresholds in the validation set. Given the clinical importance of minimizing missed high-risk cases in gastric cancer, recall was prioritized during threshold selection. Accuracy, precision, F1 score, specificity and decision-curve–based net benefit were also jointly considered to identify a threshold that balanced early detection of high-risk patients with acceptable false-positive rates.

### Machine Learning Models

Six machine learning models were developed and compared for predicting the risk of skeletal muscle loss during treatment in patients with gastric cancer, including Random Forest (RF), Extreme Gradient Boosting (XGBoost), Light Gradient Boosting Machine (LightGBM), Categorical Boosting (CatBoost), Support Vector Classifier (SVC), and Multilayer Perceptron (MLP). The candidate variables were prespecified based on prior literature, clinical relevance, and data availability. Given the moderate number of clinically relevant variables and the observed decline in predictive performance after exploratory feature selection, no additional data-driven feature selection procedure was applied before final model development. The entire set was randomly divided into a training set and a validation set at a ratio of 7:3. Hyperparameter tuning was performed in the training set using random search with five-fold cross-validation, and model generalizability was subsequently evaluated in the validation set.

Model performance was assessed in terms of discrimination, calibration, and clinical utility. Discriminative performance was evaluated using the area under the receiver operating characteristic curve (ROC-AUC), area under the precision–recall curve (PR-AUC), sensitivity, specificity, precision, and F1 score. Calibration was assessed using calibration curves, and clinical utility was evaluated by decision curve analysis (DCA).

In addition, Shapley additive explanations (SHAP) were applied to interpret the best-performing model by quantifying global feature importance and explaining individual predictions. Model development was conducted in Python using the scikit-learn library, and SHAP analysis was implemented with shap version 0.40.0.

This study was conducted and reported in accordance with the TRIPOD guidelines.

### Statistical analysis

Baseline characteristics were compared between the training and validation sets. Continuous variables were analyzed using Student’s t-test or the Mann–Whitney U test, as appropriate, and categorical variables were compared using the χ² test or Fisher’s exact test. As these comparisons were mainly descriptive, no formal adjustment for multiple testing was performed, and the resulting P values were interpreted with caution. Longitudinal changes were assessed using the Wilcoxon signed-rank test. A two-sided P value < 0.05 was considered statistically significant. Statistical analyses were conducted using Python (version 3.13).

The study’s schematic workflow was presented in Fig 1.

**Fig 1.**
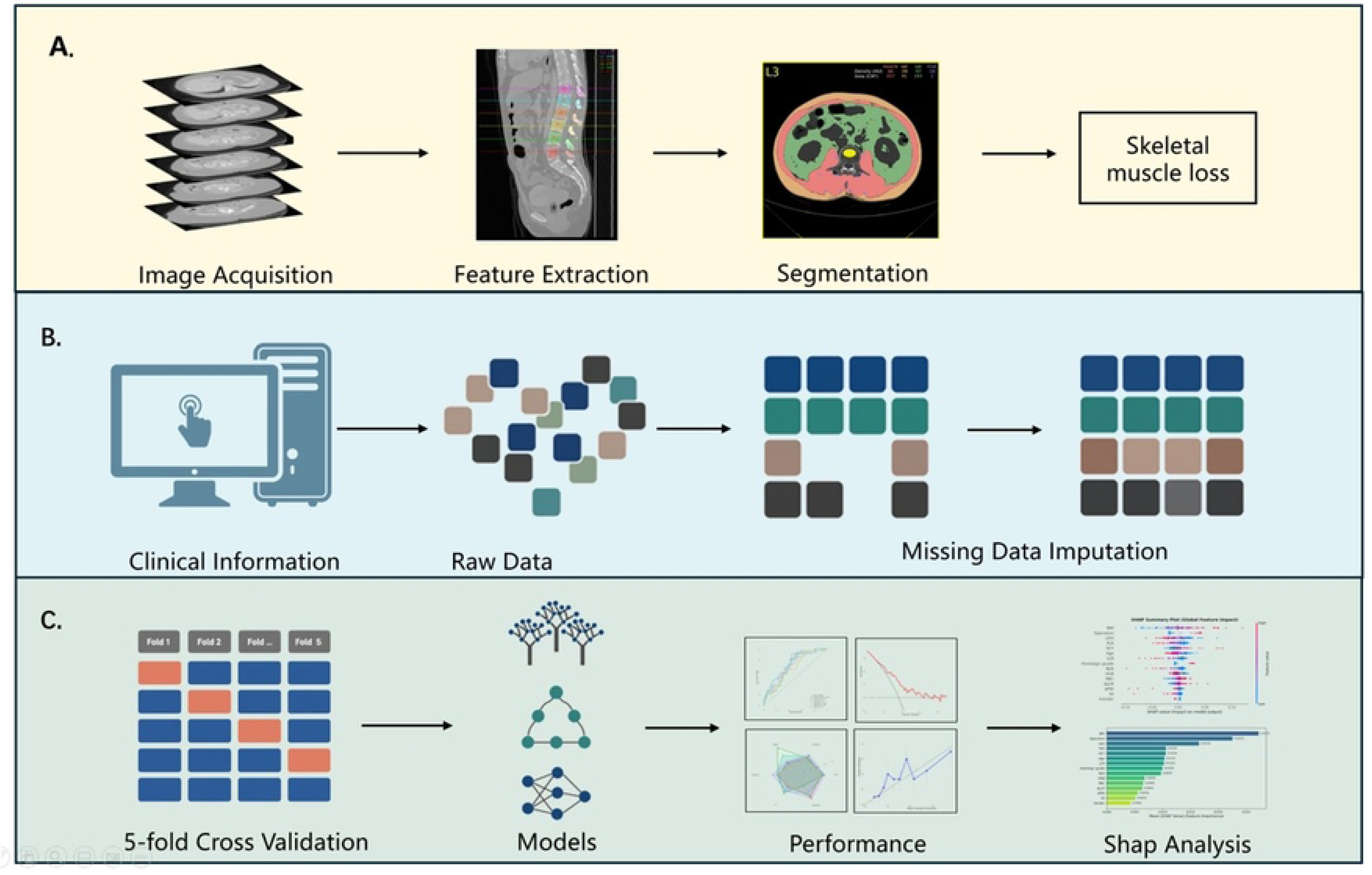
The overall workflow of the model.

## Results

### Basic characteristics

A total of 292 patients with gastric cancer were included in this study, of whom 204 were assigned to the training set and 88 to the validation set. The baseline characteristics of the study population are summarized in Table 1. Overall, the two sets were well balanced, with no significant differences in demographic characteristics, tumor-related features, or laboratory variables between the training and validation sets (all P > 0.05). Specifically, sex, age, BMI, histologic grade, tumor location, operation type, T stage, and N stage were comparable between the two groups. In addition, baseline laboratory and inflammatory/nutritional variables, including RBC, HGB, LDH, HCY, CEA, CA199, CA724, NLR, LLR, PLR, SII, PNI, and their longitudinal change features, also showed no significant between-group differences (all P > 0.05).

**Table 1.**
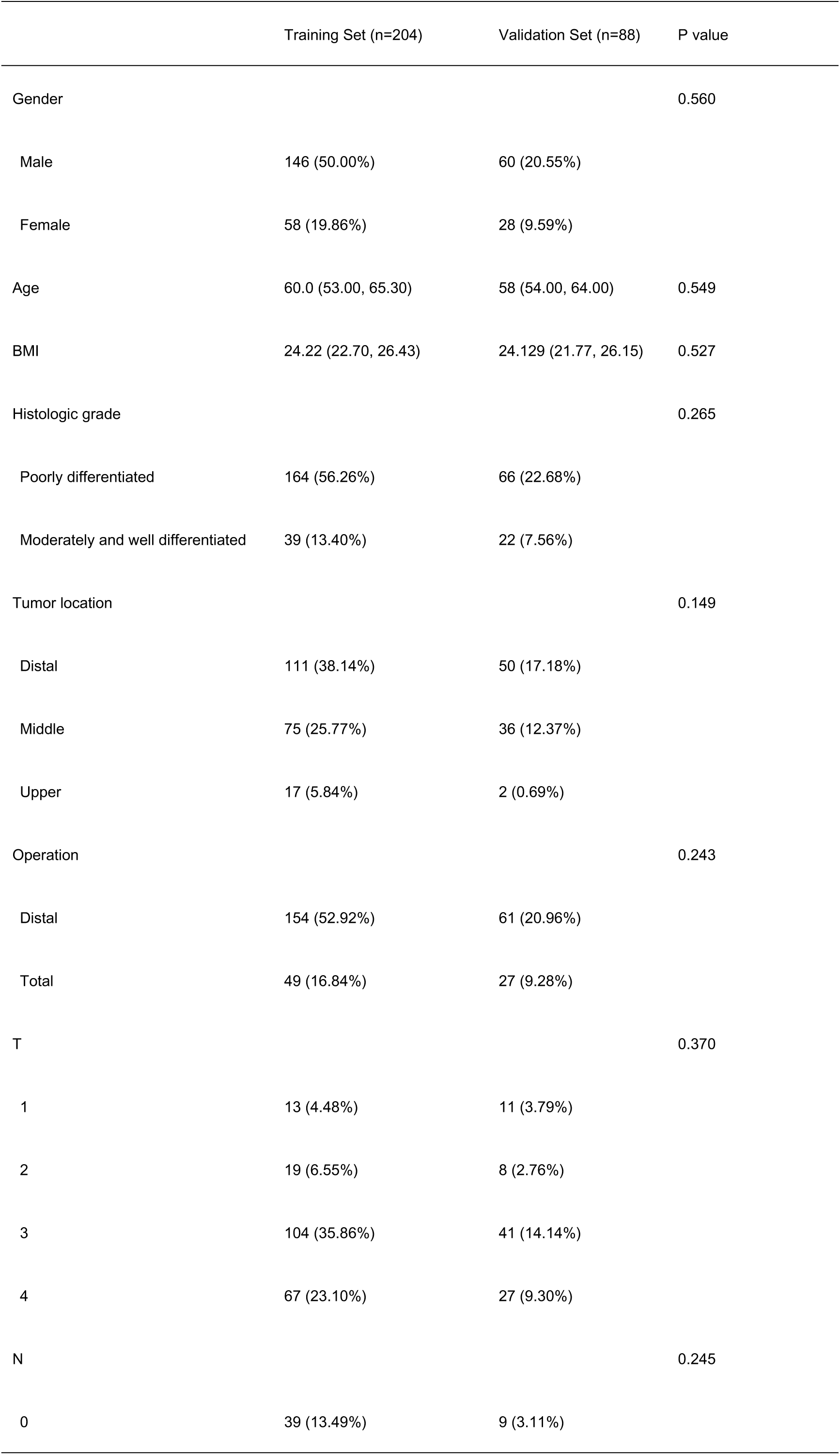
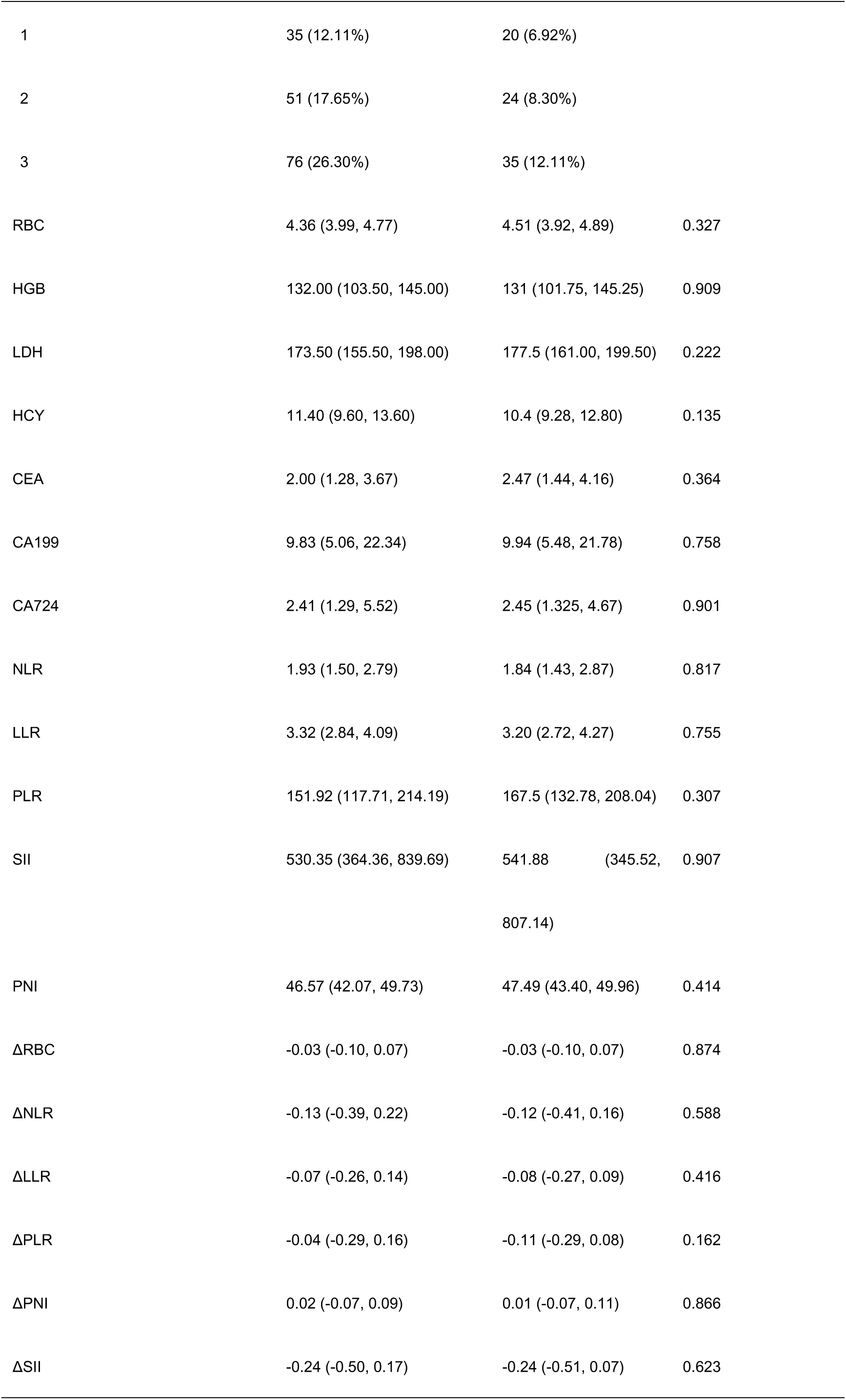
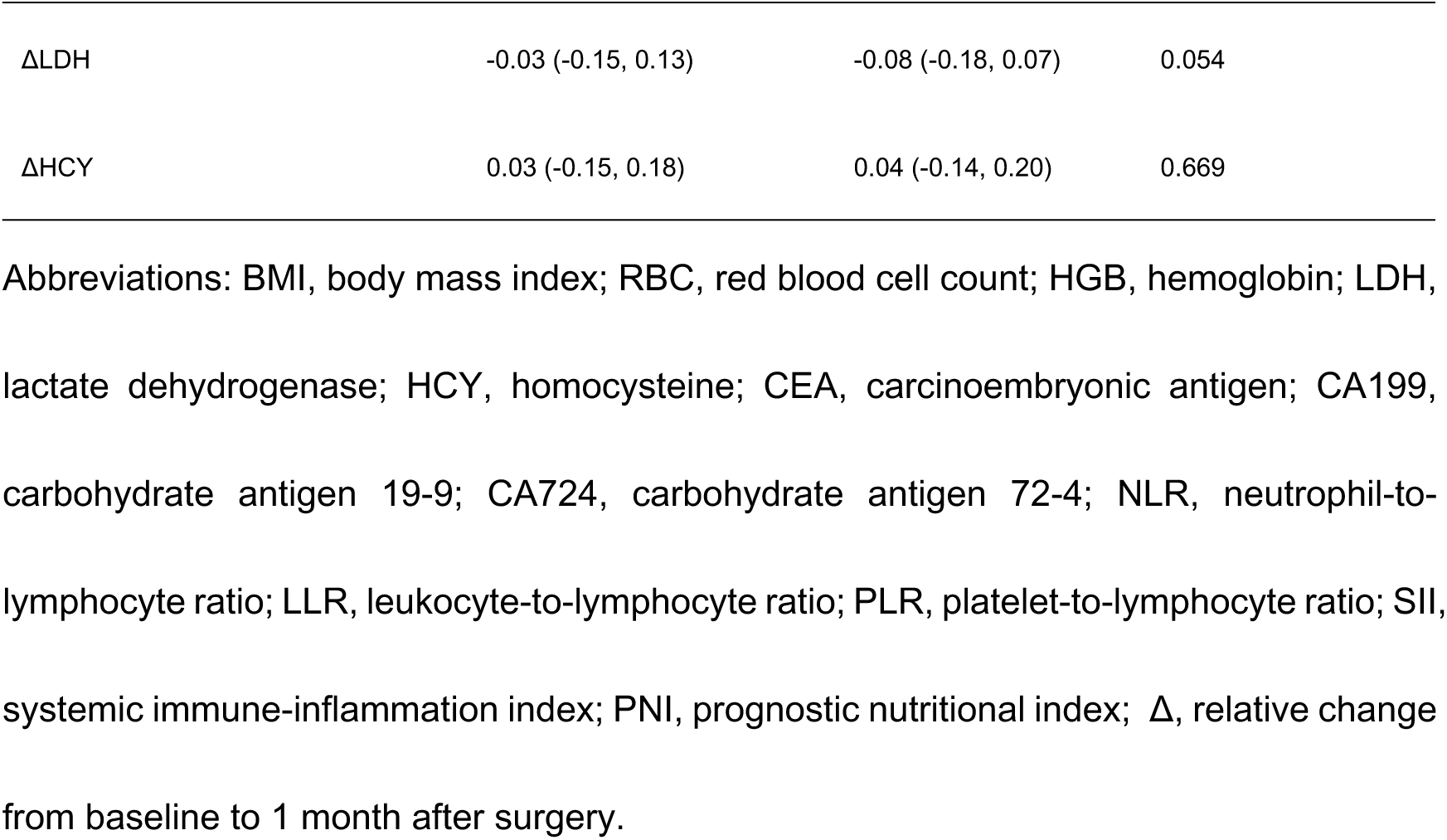
Baseline patient characteristics.

| Training Set (n=204) | Validation Set (n=88) | P value |
| --- | --- | --- |

|  |  |  |  |
| --- | --- | --- | --- |
| Gender |  |  | 0.560 |
| Male | 146 (50.00%) | 60 (20.55%) |  |
| Female | 58 (19.86%) | 28 (9.59%) |  |
| Age | 60.0 (53.00, 65.30) | 58 (54.00, 64.00) | 0.549 |
| BMI | 24.22 (22.70, 26.43) | 24.129 (21.77, 26.15) | 0.527 |
| Histologic grade |  |  | 0.265 |
| Poorly differentiated | 164 (56.26%) | 66 (22.68%) |  |
| Moderately and well differentiated | 39 (13.40%) | 22 (7.56%) |  |
| Tumor location |  |  | 0.149 |
| Distal | 111 (38.14%) | 50 (17.18%) |  |
| Middle | 75 (25.77%) | 36 (12.37%) |  |
| Upper | 17 (5.84%) | 2 (0.69%) |  |
| Operation |  |  | 0.243 |
| Distal | 154 (52.92%) | 61 (20.96%) |  |
| Total | 49 (16.84%) | 27 (9.28%) |  |
| T |  |  | 0.370 |
| 1 | 13 (4.48%) | 11 (3.79%) |  |
| 2 | 19 (6.55%) | 8 (2.76%) |  |
| 3 | 104 (35.86%) | 41 (14.14%) |  |
| 4 | 67 (23.10%) | 27 (9.30%) |  |
| N |  |  | 0.245 |
| 0 | 39 (13.49%) | 9 (3.11%) |  |
| 1 | 35 (12.11%) | 20 (6.92%) |  |
| 2 | 51 (17.65%) | 24 (8.30%) |  |
| 3 | 76 (26.30%) | 35 (12.11%) |  |
| RBC | 4.36 (3.99, 4.77) | 4.51 (3.92, 4.89) | 0.327 |
| HGB | 132.00 (103.50, 145.00) | 131 (101.75, 145.25) | 0.909 |
| LDH | 173.50 (155.50, 198.00) | 177.5 (161.00, 199.50) | 0.222 |
| HCY | 11.40 (9.60, 13.60) | 10.4 (9.28, 12.80) | 0.135 |
| CEA | 2.00 (1.28, 3.67) | 2.47 (1.44, 4.16) | 0.364 |
| CA199 | 9.83 (5.06, 22.34) | 9.94 (5.48, 21.78) | 0.758 |
| CA724 | 2.41 (1.29, 5.52) | 2.45 (1.325, 4.67) | 0.901 |
| NLR | 1.93 (1.50, 2.79) | 1.84 (1.43, 2.87) | 0.817 |
| LLR | 3.32 (2.84, 4.09) | 3.20 (2.72, 4.27) | 0.755 |
| PLR | 151.92 (117.71, 214.19) | 167.5 (132.78, 208.04) | 0.307 |
| SII | 530.35 (364.36, 839.69) | 541.88 (345.52, 807.14) | 0.907 |
| PNI | 46.57 (42.07, 49.73) | 47.49 (43.40, 49.96) | 0.414 |
| ΔRBC | -0.03 (-0.10, 0.07) | -0.03 (-0.10, 0.07) | 0.874 |
| ΔNLR | -0.13 (-0.39, 0.22) | -0.12 (-0.41, 0.16) | 0.588 |
| ΔLLR | -0.07 (-0.26, 0.14) | -0.08 (-0.27, 0.09) | 0.416 |
| ΔPLR | -0.04 (-0.29, 0.16) | -0.11 (-0.29, 0.08) | 0.162 |
| ΔPNI | 0.02 (-0.07, 0.09) | 0.01 (-0.07, 0.11) | 0.866 |
| ΔSII | -0.24 (-0.50, 0.17) | -0.24 (-0.51, 0.07) | 0.623 |
| ΔLDH | -0.03 (-0.15, 0.13) | -0.08 (-0.18, 0.07) | 0.054 |
| ΔHCY | 0.03 (-0.15, 0.18) | 0.04 (-0.14, 0.20) | 0.669 |

Abbreviations: BMI, body mass index; RBC, red blood cell count; HGB, hemoglobin; LDH, lactate dehydrogenase; HCY, homocysteine; CEA, carcinoembryonic antigen; CA199, carbohydrate antigen 19-9; CA724, carbohydrate antigen 72-4; NLR, neutrophil-to-lymphocyte ratio; LLR, leukocyte-to-lymphocyte ratio; PLR, platelet-to-lymphocyte ratio; SII, systemic immune-inflammation index; PNI, prognostic nutritional index; $\Delta$ , relative change from baseline to 1 month after surgery.

### Threshold selection

To identify a clinically meaningful operating point, model performance was systematically evaluated across a range of decision thresholds in the validation set. In this clinical context, false-positive classifications were considered more acceptable than false-negative classifications, because missing patients truly at high risk of skeletal muscle loss could delay timely nutritional and supportive interventions. Therefore, greater emphasis was placed on maintaining high recall while also considering overall classification balance.

At lower thresholds (0.25–0.40), the MLP model achieved very high recall (0.9583–1.0000), but specificity remained low (0–0.25), indicating limited ability to correctly identify low-risk patients. In contrast, higher thresholds (0.50–0.60) substantially improved specificity (0.90–0.975) at the cost of a marked reduction in recall (0.0208–0.4375). Threshold 0.45 was selected as the final operating point because it provided the most balanced overall performance. At this threshold, the MLP model achieved the highest accuracy (0.6932) and F1 score (0.7477), while maintaining a relatively high recall (0.8333) and a moderate specificity (0.5250). The corresponding precision was 0.6780, and the net benefit was 0.2779. Taken together, these findings indicate that a threshold of 0.45 represented the most clinically appropriate compromise between minimizing missed high-risk cases and limiting excessive false-positive classifications (Table 2).

**Table 2.**
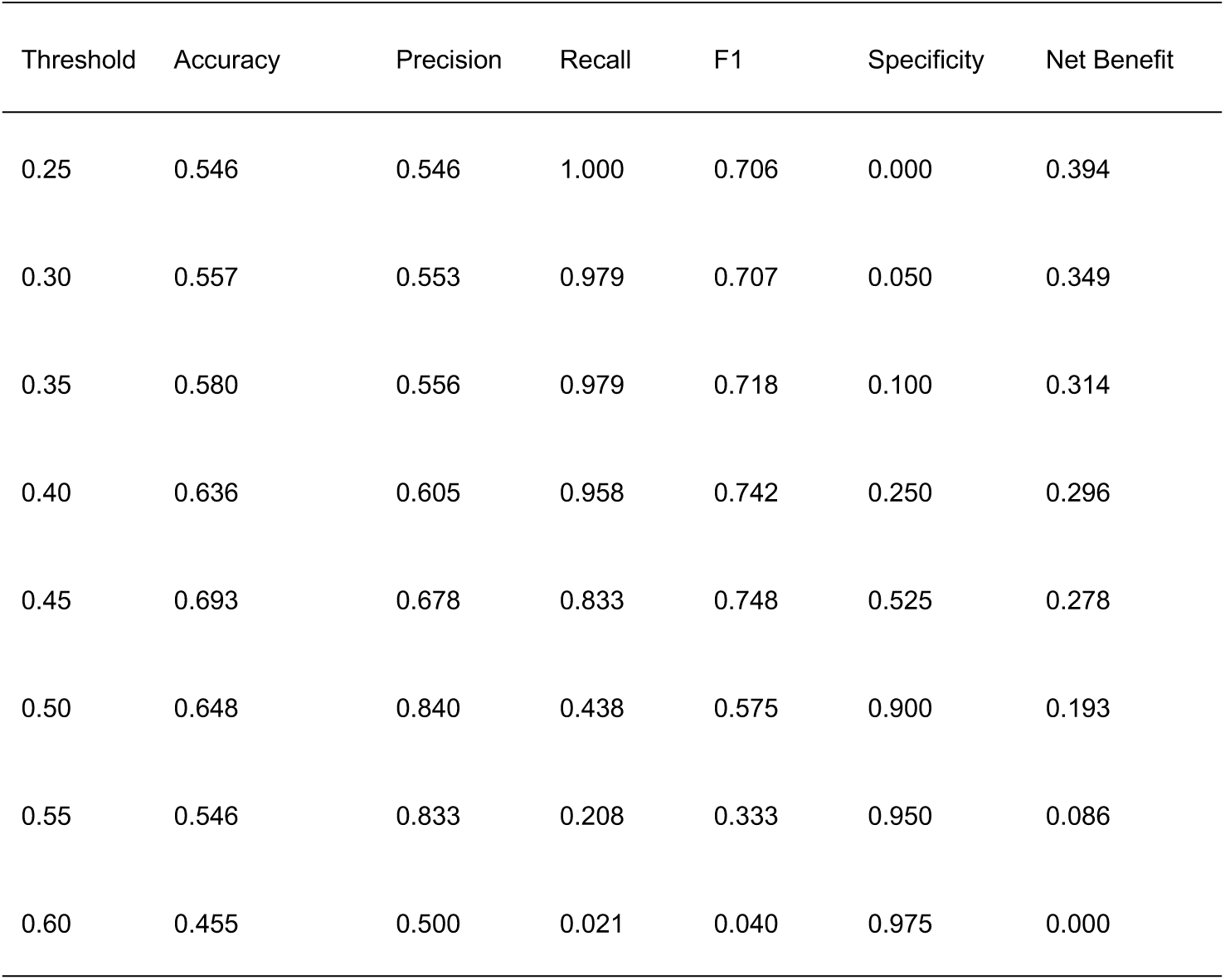
Threshold-specific performance of the MLP model in the validation set.

### Performance of different models

The predictive performance of the six machine learning models for significant skeletal muscle loss in the validation set is summarized in Table 3 and Fig 2. Among all evaluated models, the MLP model achieved the best overall discriminative performance, with the highest AUC of 0.757, followed by SVC (0.668), LightGBM (0.649), Random Forest (0.633), XGBoost (0.597), and CatBoost (0.555). Precision–recall analysis showed a similar pattern, with MLP again yielding the highest PR-AUC (0.745), followed by SVC (0.677), LightGBM (0.640), Random Forest (0.626), XGBoost (0.599), and CatBoost (0.578).

**Fig 2.**
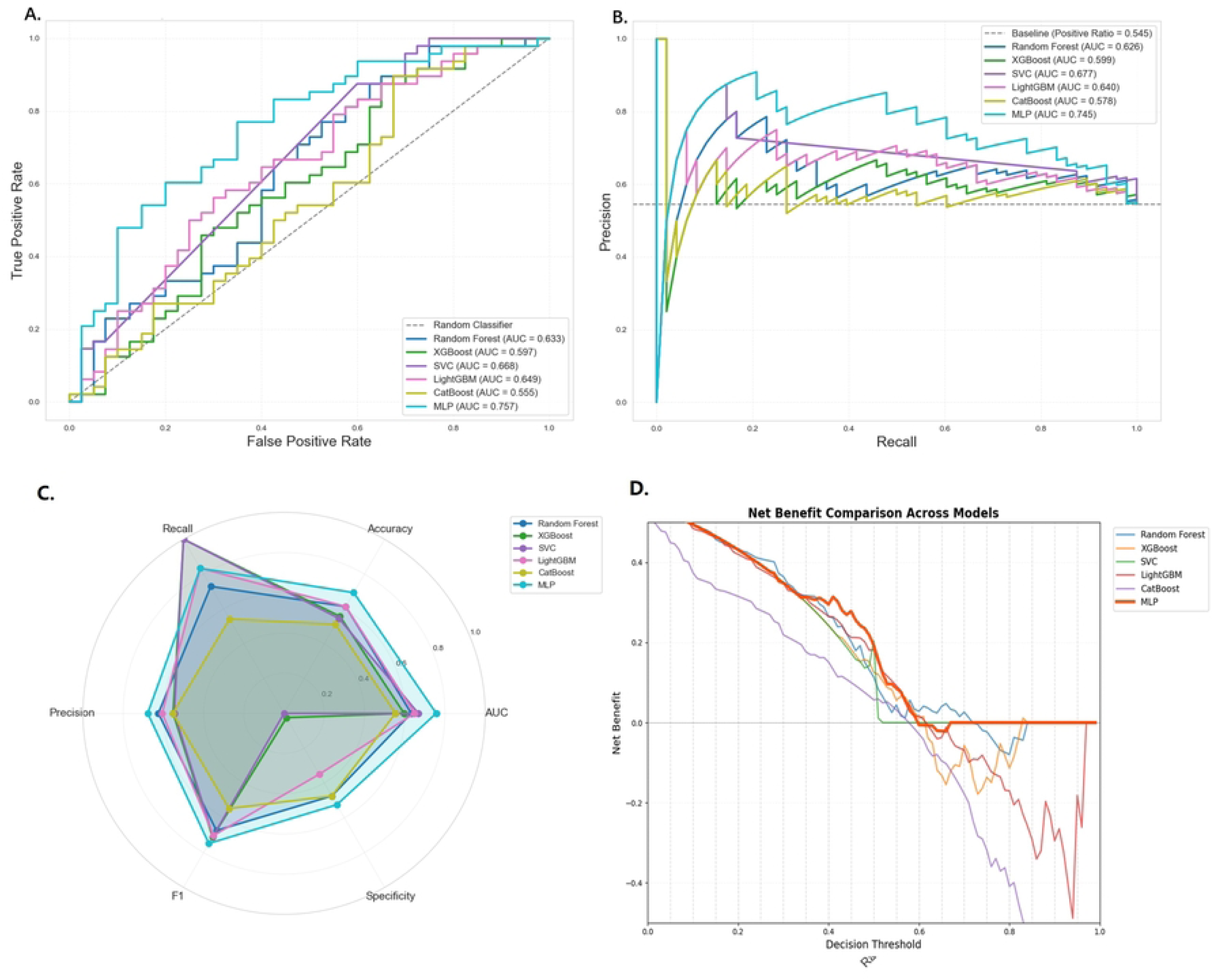
Assessment of model classification performance. (A) Receiver operating characteristic (ROC) curves of the six models. (B) Precision–recall (PR) curves of the six models. The dashed line indicates the baseline positive rate. (C) Radar plot comparing the main performance metrics of each model, including AUC, accuracy, recall, precision, F1 score, and specificity. (D) Decision curve analysis (DCA) showing the net benefit of each model across a range of decision thresholds.

**Table 3.**
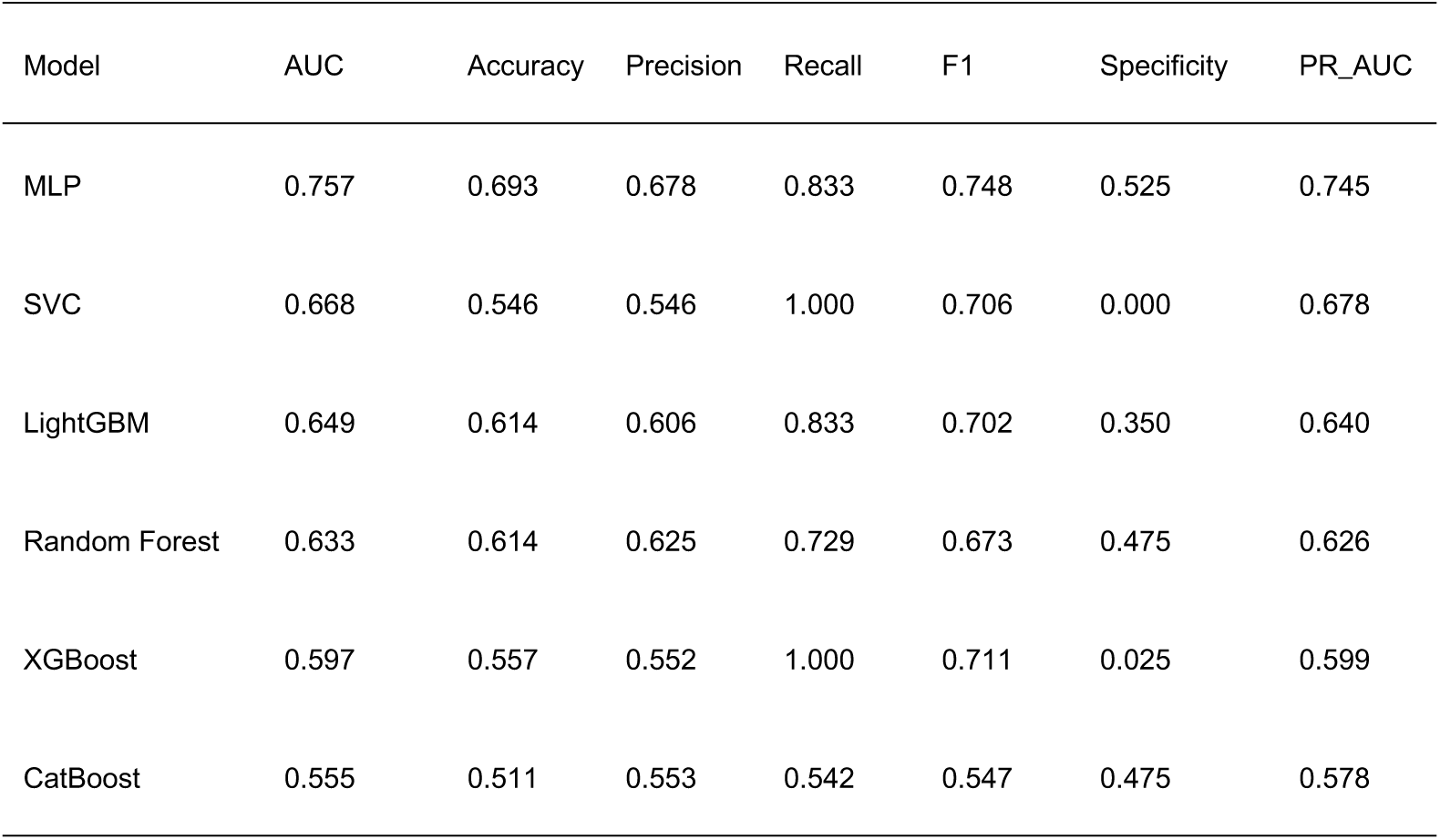
Performance comparison of models at the decision threshold of 0.45 in the validation set.

In addition to discrimination, MLP also showed the most balanced classification performance in the validation set, with an accuracy of 0.693, precision of 0.678, recall of 0.833, F1 score of 0.748, and specificity of 0.525. Although SVC and XGBoost achieved a recall of 1.000, both models showed very low specificity (0.000 and 0.025, respectively), indicating limited ability to correctly identify patients at low risk of skeletal muscle loss. LightGBM and Random Forest showed moderate overall performance, whereas CatBoost demonstrated the weakest discrimination and the lowest overall classification performance. Taken together, these findings indicate that MLP provided the most favorable balance between sensitivity and specificity for predicting subsequent skeletal muscle loss.

Decision curve analysis further demonstrated that the MLP model provided the greatest net benefit across a relatively broad range of threshold probabilities, supporting its potential clinical utility (Figs 2D and 3A-F). Calibration curves indicated acceptable agreement between predicted and observed probabilities, and the corresponding Brier score also suggested reasonable calibration performance of the MLP model (Fig 3G-L). Therefore, considering discriminative ability, classification performance, calibration, and clinical usefulness together, the MLP model was selected as the final model for subsequent interpretation.

**Fig 3.**
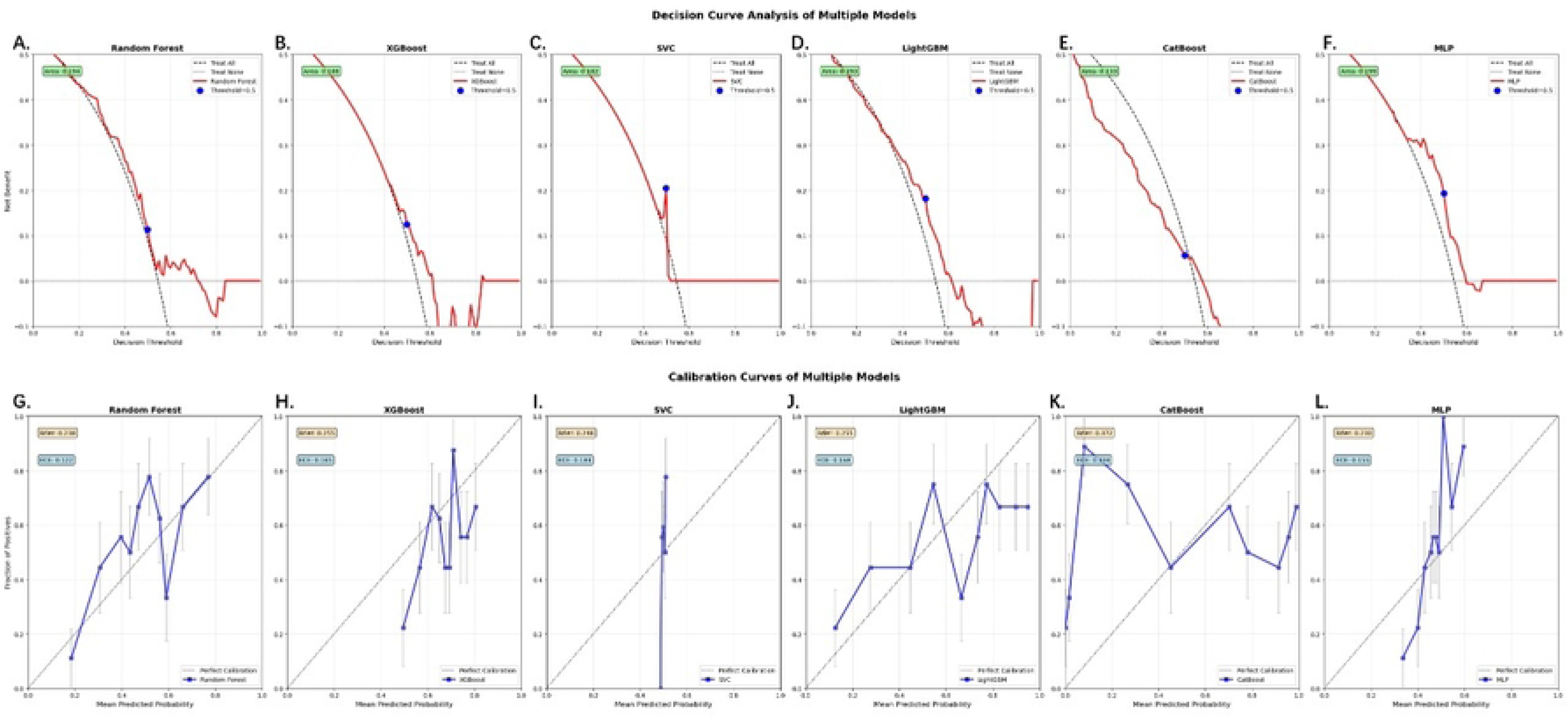
Decision curve analysis and calibration curves of the six machine learning models in the validation set. (A–F) Decision curve analysis of Random Forest, XGBoost, SVC, LightGBM, CatBoost, and MLP, respectively. Blue dots indicate the selected threshold of 0.45. (G–L) Calibration curves of the corresponding models. The gray diagonal line indicates perfect calibration. The Brier score and expected calibration error (ECE) for each model are also shown within the corresponding calibration panels.

We further compared the predictive performance of the MLP model with and without longitudinal dynamic features in Table 4. Overall, incorporation of dynamic features improved model performance. Specifically, the MLP model with dynamic features achieved a higher AUC (0.757 vs. 0.709), accuracy (0.693 vs. 0.591), recall (0.833 vs. 0.708), F1 score (0.748 vs. 0.654), and specificity (0.525 vs. 0.450) than the model using baseline features alone. Precision was also modestly improved (0.678 vs. 0.607). Although the PR-AUC of the model with dynamic features was slightly lower than that of the baseline-only model (0.745 vs. 0.759), the overall results indicate that longitudinal dynamic features provided additional predictive information and improved the overall discriminative and classification performance of the MLP model.

**Table 4.** Comparison of predictive performance of the MLP model with and without longitudinal dynamic features.

| Model version | AUC | Accuracy | Precision | Recall | F1 | Specificity | PR_AUC |
| --- | --- | --- | --- | --- | --- | --- | --- |
| With dynamic features | 0.757 | 0.693 | 0.678 | 0.833 | 0.748 | 0.525 | 0.745 |
| Without dynamic features | 0.709 | 0.591 | 0.607 | 0.708 | 0.654 | 0.450 | 0.759 |

The SHAP analysis of the final MLP model is presented in Fig 4. Among the evaluated variables, BMI and operation type were identified as the two most influential features, followed by LDH, PLR, HCY, age, and LLR. Other variables with notable contributions included histologic grade, NLR, HGB, RBC, ΔLLR, ΔPNI, SII, and gender. Overall, the SHAP results suggested that the prediction of skeletal muscle loss was jointly influenced by baseline physiological reserve, treatment-related burden, and inflammatory/metabolic status.

**Fig 4.**
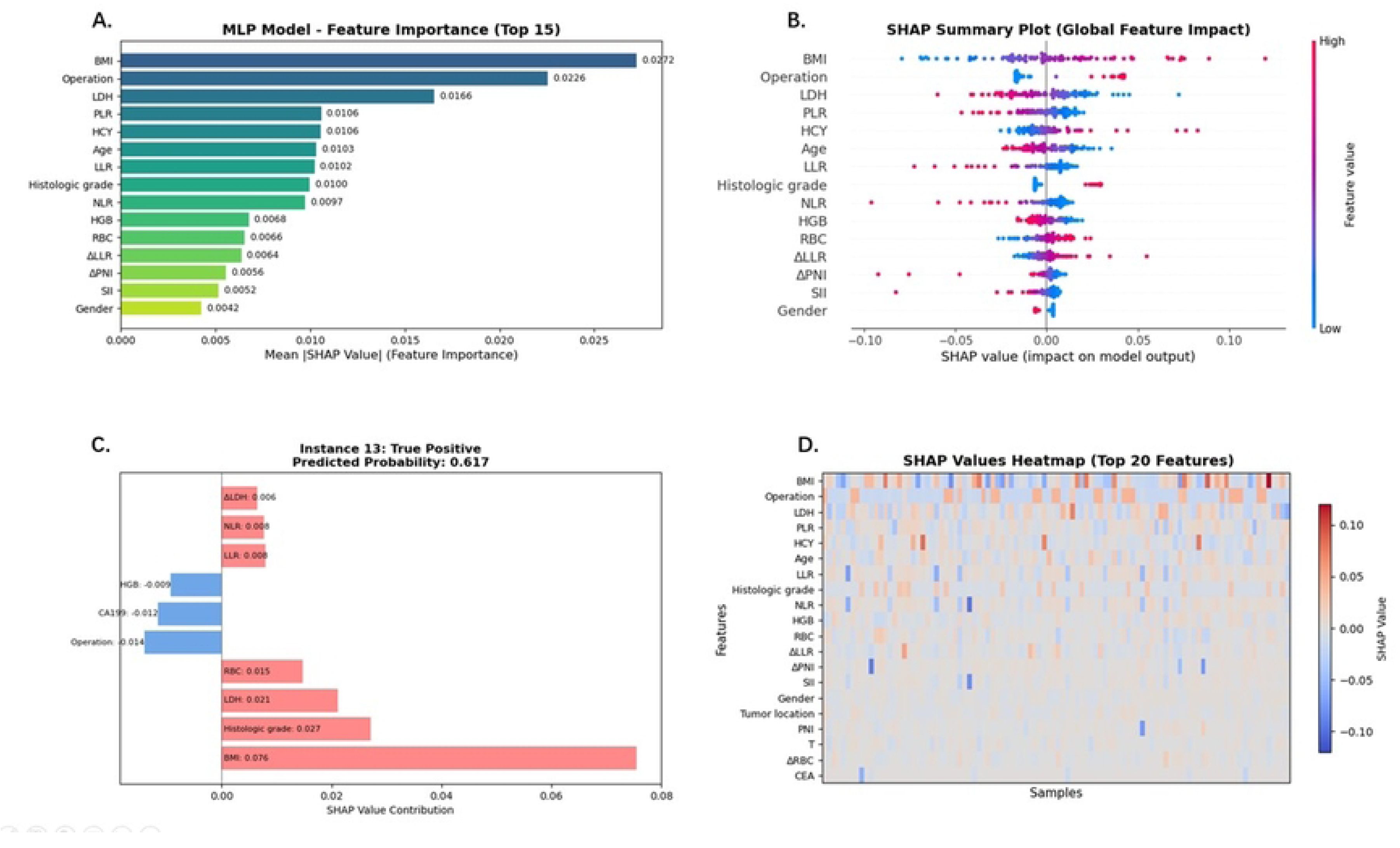
SHAP interpretability analysis of the best prediction model. (A) Global feature importance ranked by mean absolute SHAP value. (B) SHAP summary plot showing the distribution of feature effects on model output. Each point represents one patient, and color indicates the feature value from low to high. (C) Individual SHAP force plot for a representative true-positive case, illustrating the contribution of each feature to the predicted probability. (D) Heatmap of SHAP values for the top 20 features across all patients.

## Discussion

In the present study, we developed and compared six machine learning models using routinely available clinical data to enable early prediction of subsequent significant skeletal muscle loss in patients with gastric cancer undergoing radical gastrectomy followed by adjuvant chemotherapy. Among the evaluated models, the MLP model showed the best overall performance in the validation set. These findings suggest that preoperative baseline variables together with early postoperative dynamic changes, assessed at 1 month after surgery, already contain clinically meaningful signals associated with later skeletal muscle loss. Therefore, early risk stratification before overt muscle depletion becomes evident on follow-up CT appears feasible and may provide a basis for earlier supportive interventions during treatment.

The main clinical implication of this study is the forward shift of the prediction window. CT-based body composition assessment remains the standard approach for quantifying skeletal muscle status, but in routine practice it is primarily used for objective measurement and retrospective confirmation rather than for early warning. In contrast, although skeletal muscle loss in our study was defined by standardized CT-based changes in SMI, the predictive model itself relied mainly on clinical data that are routinely available before surgery and 1 month after surgery. This design is more closely aligned with real-world clinical management. From a clinical perspective, the present model should be considered an early risk stratification tool rather than a replacement for CT-based assessment. Patients identified as high risk may benefit from earlier nutritional support, exercise-based rehabilitation, metabolic management, and closer follow-up, with the goal of reducing subsequent clinically relevant skeletal muscle loss[7, 18].

Compared with previous studies, our work places greater emphasis on clinical accessibility rather than maximal imaging input. Hsu et al. developed an explainable machine learning model in ovarian cancer using clinical variables and their longitudinal changes to predict skeletal muscle loss during surgery and adjuvant chemotherapy, demonstrating that non-imaging clinical information can provide meaningful predictive value[10]. By contrast, Huang et al. developed a CT-based deep learning model for prognosis prediction in patients with esophageal squamous cell carcinoma receiving immunotherapy combined with chemotherapy, highlighting the value of imaging-derived deep features in outcome prediction[19]. Taken together, these studies represent two complementary directions in this field: imaging-centered artificial intelligence models and clinically deployable models based on routinely available variables. Our study is more consistent with the latter approach, as CT-derived body composition was used only for outcome definition rather than as a required input for the final recommended model. In addition, our fixed time-anchored design—baseline assessment before surgery, early postoperative clinical evaluation at 1 month, and follow-up CT at 3 months after initiation of adjuvant chemotherapy—better reflects the actual treatment trajectory of patients with gastric cancer.

The explainability analysis further improved the clinical interpretability of the model. Overall, the most influential features could be broadly grouped into baseline nutritional and physiological reserve, operation-related burden, and inflammatory/metabolic status[12]. This pattern suggests that skeletal muscle loss in this setting is unlikely to be explained by nutritional insufficiency alone, but rather reflects the combined effects of surgical stress, recovery capacity, systemic inflammation, and metabolic disturbance. Notably, operation type had relatively high importance in the model. In addition to differences in surgical trauma and postoperative recovery burden, total gastrectomy generally involves more extensive tissue resection than distal gastrectomy, including removal of omental visceral fat, which may partly affect postoperative energy reserve and metabolic status and thereby contribute to later skeletal muscle loss[20]. From this perspective, skeletal muscle loss may be better understood as a multidimensional marker of peri-treatment physiological imbalance rather than as an isolated nutritional endpoint[18].

This study has several strengths. First, the model was developed using routinely available clinical variables and their early dynamic changes rather than repeated body composition measurements, which enhances its potential clinical applicability. Second, the study design incorporated clearly defined time points and therefore captured a relatively complete clinical trajectory from the preoperative period to early postoperative recovery and adjuvant treatment. Third, CT-based body composition analysis was performed using the automated Comp2Comp platform with radiologist review for quality control, thereby improving the objectivity and standardization of outcome assessment.

Several limitations should also be acknowledged. First, this was a single-center retrospective study, and the generalizability of the model still requires external validation in larger multicenter sets. Second, because the model used preoperative and 1-month postoperative clinical data, the predictive signals captured by the model likely reflect not only adjuvant treatment-related effects but also the impact of surgical stress and postoperative recovery. Accordingly, the outcome predicted in this study may be more accurately interpreted as the risk of subsequent skeletal muscle loss during the perioperative and early adjuvant-treatment period, rather than chemotherapy-related muscle loss alone. Third, although we defined significant skeletal muscle loss as an SMI decrease of at least 5%, consistent with previous studies, some heterogeneity in threshold definitions across studies should be recognized. In addition, the threshold was selected and evaluated within the same validation set, which may have resulted in optimistic performance estimates.

In conclusion, we developed an interpretable machine learning model based on routinely available preoperative and early postoperative clinical data for early prediction of significant skeletal muscle loss in patients with gastric cancer after radical gastrectomy and adjuvant chemotherapy. The MLP model demonstrated the best overall performance. This framework provides a clinically relevant strategy for early identification of patients at risk of skeletal muscle loss and may facilitate more individualized nutritional support and supportive care during treatment.

## Acknowledgments

The authors would like to thank all participating patients.

## Ethics approval and consent to participate

This study strictly adhered to the principles outlined in the Helsinki Declaration and received formal approval from the Ethics Committee of Yantai Yuhuangding Hospital, with the approval number being 2025-115.

Informed consent was waived by Ethics Committee of Yantai Yuhuangding Hospital due to the retrospective design and use of fully anonymized patient data. Registry and the Registration No. of the study/trial: N/A

Animal Studies: N/A

## Author Contributions

Hanzheng Wang: Conceptualization, Formal Analysis, Investigation, Writing – Original Draft, Visualization, Project Administration.

Kun Ma: Conceptualization, Writing – Review & Editing, Methodology.

Jiezhi Lin: Conceptualization, Writing – Review & Editing, Writing – Review & Editing, Supervision.

Juntao Zhu: Conceptualization, Writing – Review & Editing, Supervision.

Mengchen Sun: Validation, Formal Analysis, Investigation, Resources, Data Curation, Writing – Original Draft, Visualization.

Shubin Liang: Software, Investigation, Resources.

Bin Yang: Conceptualization, Methodology, Validation, Formal Analysis, Writing – Review & Editing, Visualization, Writing – Original Draft.

Linsong Mu: Resources, Writing – Review & Editing, Data Curation.

Zhongchuan Lv: Writing – Original Draft, Writing – Review & Editing, Visualization, Funding Acquisition.

All authors have read and approved the final version of the manuscript.

## Data availability statement

The de-identified minimal dataset underlying the findings of this study is publicly available in the GitHub repository at: https://github.com/HzWang124144/gastric-cancer-skeletal-muscle-loss/

## Declaration of generative AI and AI-assisted technologies in the writing process

During the preparation of this work, the authors used ChatGPT (OpenAI) to assist with language editing, text organization, and improving the clarity of the manuscript. After using this tool, the authors reviewed and edited the content as needed and take full responsibility for the content of the publication.

## Reference

1. Bray F, Laversanne M, Sung H, Ferlay J, Siegel RL, Soerjomataram I, et al. Global cancer statistics 2022: GLOBOCAN estimates of incidence and mortality worldwide for 36 cancers in 185 countries. CA Cancer J Clin. 2024;74(3):229–63. Epub 20240404. doi: 10.3322/caac.21834. PubMed PMID: 38572751.

2. Kim IH, Kang SJ, Choi W, Seo AN, Eom BW, Kang B, et al. Korean Practice Guidelines for Gastric Cancer 2024: An Evidence-based, Multidisciplinary Approach (Update of 2022 Guideline). J Gastric Cancer. 2025;25(1):5–114. doi: 10.5230/jgc.2025.25.e11. PubMed PMID: 39822170; PubMed Central PMCID: PMCPMC11739648.

3. Bossi P, Delrio P, Mascheroni A, Zanetti M. The Spectrum of Malnutrition/Cachexia/Sarcopenia in Oncology According to Different Cancer Types and Settings: A Narrative Review. Nutrients. 2021;13(6). Epub 20210609. doi: 10.3390/nu13061980. PubMed PMID: 34207529; PubMed Central PMCID: PMCPMC8226689.

4. Jogiat UM, Sasewich H, Turner SR, Baracos V, Eurich DT, Filafilo H, et al. Sarcopenia Determined by Skeletal Muscle Index Predicts Overall Survival, Disease-free Survival, and Postoperative Complications in Resectable Esophageal Cancer: A Systematic Review and Meta-analysis. Ann Surg. 2022;276(5):e311–e8. Epub 20220706. doi: 10.1097/sla.0000000000005452. PubMed PMID: 35794004.

5. Li X, Ding P, Wu J, Wu H, Yang P, Guo H, et al. Preoperative sarcopenia and postoperative accelerated muscle loss negatively impact survival after resection of locally advanced gastric cancer. BMC Cancer. 2025;25(1):269. Epub 20250214. doi: 10.1186/s12885-025-13674-3. PubMed PMID: 39953409; PubMed Central PMCID: PMCPMC11829415.

6. Brown LR, Sousa MS, Yule MS, Baracos VE, McMillan DC, Arends J, et al. Body weight and composition endpoints in cancer cachexia clinical trials: Systematic Review 4 of the cachexia endpoints series. J Cachexia Sarcopenia Muscle. 2024;15(3):816–52. Epub 20240513. doi: 10.1002/jcsm.13478. PubMed PMID: 38738581; PubMed Central PMCID: PMCPMC11154800.

7. Jiang Y, Zhao Y, Dai J, Yang Q, Tang X, Fu L, et al. Imaging Cancer-associated Cachexia: Utilizing Clinical Imaging Modalities for Early Diagnosis. Radiol Imaging Cancer. 2025;7(4):e240291. doi: 10.1148/rycan.240291. PubMed PMID: 40476859; PubMed Central PMCID: PMCPMC12304533.

8. Rinninella E, Cintoni M, Raoul P, Pozzo C, Strippoli A, Bria E, et al. Muscle mass, assessed at diagnosis by L3-CT scan as a prognostic marker of clinical outcomes in patients with gastric cancer: A systematic review and meta-analysis. Clin Nutr. 2020;39(7):2045–54. Epub 20191101. doi: 10.1016/j.clnu.2019.10.021. PubMed PMID: 31718876.

9. Zhan C, Bu J, Li S, Huang X, Quan Z. Postoperative skeletal muscle loss as a prognostic indicator of clinical outcomes in patients with gastric cancer: a systematic review and meta-analysis. J Gastrointest Surg. 2025;29(2):101898. Epub 20241126. doi: 10.1016/j.gassur.2024.101898. PubMed PMID: 39608746.

10. Hsu WH, Ko AT, Weng CS, Chang CL, Jan YT, Lin JB, et al. Explainable machine learning model for predicting skeletal muscle loss during surgery and adjuvant chemotherapy in ovarian cancer. J Cachexia Sarcopenia Muscle. 2023;14(5):2044–53. Epub 20230712. doi: 10.1002/jcsm.13282. PubMed PMID: 37435785; PubMed Central PMCID: PMCPMC10570082.

11. Chen Y, Liu C, Zheng X, Liu T, Xie H, Lin SQ, et al. Machine learning to identify precachexia and cachexia: a multicenter, retrospective cohort study. Support Care Cancer. 2024;32(10):630. Epub 20240903. doi: 10.1007/s00520-024-08833-4. PubMed PMID: 39225814; PubMed Central PMCID: PMCPMC11371878.

12. Geppert J, Rohm M. Cancer cachexia: biomarkers and the influence of age. Mol Oncol. 2024;18(9):2070–86. Epub 20240227. doi: 10.1002/1878-0261.13590. PubMed PMID: 38414161; PubMed Central PMCID: PMCPMC11467804.

13. Lipshitz M, Visser J, Anderson R, Nel DG, Smit T, Steel HC, et al. Relationships of emerging biomarkers of cancer cachexia with quality of life, appetite, and cachexia. Support Care Cancer. 2024;32(6):349. Epub 20240514. doi: 10.1007/s00520-024-08549-5. PubMed PMID: 38744744; PubMed Central PMCID: PMCPMC11093781.

14. Giddings R, Joseph A, Callender T, Janes SM, van der Schaar M, Sheringham J, et al. Factors influencing clinician and patient interaction with machine learning-based risk prediction models: a systematic review. Lancet Digit Health. 2024;6(2):e131–e44. doi: 10.1016/s2589-7500(23)00241-8. PubMed PMID: 38278615.

15. Lee MW, Jeon SK, Paik WH, Yoon JH, Joo I, Lee JM, et al. Prognostic value of initial and longitudinal changes in body composition in metastatic pancreatic cancer. J Cachexia Sarcopenia Muscle. 2024;15(2):735–45. Epub 20240208. doi: 10.1002/jcsm.13437. PubMed PMID: 38332658; PubMed Central PMCID: PMCPMC10995276.

16. Blankemeier L, Desai A, Zambrano Chaves JM, Wentland A, Yao S, Reis E, et al. Comp2Comp: Open-Source Body Composition Assessment on Computed Tomography. arXiv [Internet]. 2023 2026 Apr 22; 2302.06568. Available from: https://arxiv.org/abs/2302.06568.

17. Lee JH, Hyung S, Lee J, Choi SH. Visceral adiposity and systemic inflammation in the obesity paradox in patients with unresectable or metastatic melanoma undergoing immune checkpoint inhibitor therapy: a retrospective cohort study. J Immunother Cancer. 2022;10(8). doi: 10.1136/jitc-2022-005226. PubMed PMID: 36002189; PubMed Central PMCID: PMCPMC9413167.

18. Weimann A, Bezmarevic M, Braga M, Correia M, Funk-Debleds P, Gianotti L, et al. ESPEN guideline on clinical nutrition in surgery - Update 2025. Clin Nutr. 2025;53:222–61. Epub 20250903. doi: 10.1016/j.clnu.2025.08.029. PubMed PMID: 40957230.

19. Huang X, Huang Y, Li P, Xu K. CT-Based Deep Learning Predicts Prognosis in Esophageal Squamous Cell Cancer Patients Receiving Immunotherapy Combined with Chemotherapy. Acad Radiol. 2025;32(6):3397–409. Epub 20250215. doi: 10.1016/j.acra.2025.01.046. PubMed PMID: 39956748.

20. Kang MK, Lee HJ. Impact of malnutrition and nutritional support after gastrectomy in patients with gastric cancer. Ann Gastroenterol Surg. 2024;8(4):534–52. Epub 20240316. doi: 10.1002/ags3.12788. PubMed PMID: 38957563; PubMed Central PMCID: PMCPMC11216795.

